# Incident Chronic Kidney Disease in meat-eaters, fish-eaters, and vegetarians: A population-based prospective study

**DOI:** 10.64898/2025.12.30.25343209

**Authors:** Catharina J. Candussi, William Bell, Marko Mutapcic, Alysha S. Thompson, Sabine Rohrmann, Aedín Cassidy, Tilman Kühn, Martina Gaggl

## Abstract

**Introduction:** The global prevalence of chronic kidney disease (CKD) is rising and initial studies suggest that diets predominantly based on greater inclusion of plant foods may be associated with lower CKD risk. As population-based studies are lacking, we investigated the association between habitual plant-based diets and CKD in the UK Biobank cohort.

**Methods:** The UK Biobank is a large prospective cohort study of participants aged 40-69 years. Habitual diet was assessed using a baseline food frequency questionnaire, and participants were classified into five dietary groups: high meat eaters, low meat eaters, poultry eaters, pescatarians, and vegetarians. To assess the risk of CKD across these groups, we conducted multivariable Cox proportional hazard regression analyses.

**Results:** During follow up of 12.9 years 23,084 out of 416,584 developed CKD. Compared to high meat eaters, only vegetarians had a statistically significant lower risk of CKD [HR = 0.81, 95% CI: 0.71– 0.93].

**Conclusion:** This is the first population-based study on plant-based diet types and CKD risk. Our findings suggest that vegetarian diets are associated with a lower risk of CKD. Future research is needed to assess the feasibility and acceptability of plant-based diets for the prevention of CKD and other chronic diseases.

## Introduction

Plant-based diets have received substantial attention in recent years, partly due to evidence linking high consumption of animal-based foods with adverse health outcomes.^1–3^ Moreover, plant-based diets are associated with both better cardiometabolic health and a lower environmental impact.^4,5^ The prevalence of CKD is increasing worldwide, yet evidence on the potential role of plant-based diets in this vulnerable patient population is limited. In particular, data on the role of habitual dietary patterns in CKD prevention remain scarce. Several studies using dietary indices, such as the healthy plant-based diet index, suggest that individuals with reduced kidney function may benefit from a healthy, balanced diet similar to that recommended for the general population.^6–8^ Initial evidence also indicates a lower incidence of CKD among those consuming diets rich in high-quality plant foods and low in animal products.^9^ However, these studies typically assessed diet using post hoc diet quality scores, and there is a lack of data examining habitual diet types in relation to CKD.

The aim of this study was to assess the associations between habitual diet groups —high meat eaters, low meat eaters, poultry eaters, fish-eaters, vegetarians and vegans — and incident CKD using data from the population-based UK Biobank study. Our hypothesis was that a habitual diet with a greater emphasis on plant-based foods would be associated with a lower incidence of CKD, consistent with their known cardiometabolic benefits in the general population.

## Methods

We used data from the UK Biobank, a large prospective cohort comprising over 500,000 participants aged 40–69 years at the time of recruitment. Baseline data collection was carried out between 2006 and 2010 at 22 assessment centres across Wales, Scotland and England. At baseline recruitment, participants completed a comprehensive touchscreen questionnaire that gathered information on socioeconomic status, lifestyle factors and health conditions. Dietary intake was assessed using a food frequency questionnaire (FFQ), capturing the consumption of 29 specific food items. In addition, biological material and physical measurements were collected by trained personnel. Further information on the study design, recruitment process, and baseline assessments has been documented elsewhere^10,11^ The UK Biobank study received ethical approval from the National Health Service North West Multi-Centre Research Ethics Committee. The initial approval was granted in 2011 under the reference number 11/NW/0382. Subsequent renewals have occurred every five years, with approvals granted in 2016 (16/NW/0274) and 2021 (21/NW/0157). ^12,13^ All participants provided written informed consent. For the current analysis, individuals with missing dietary frequency data at baseline (n = 7,748), missing continuous covariates (n = 68,777), and prevalent CKD (n = 9,057) were excluded from the analysis, resulting in a final cohort of 416,584 participants. Additionally, nutritional intakes of only 178,209 participants were analyzed due to incomplete dietary data on 24-hour recall (Supplementary Figure S1).

### Dietary Assessment and Categorization

Similar to Parra-Soto et al. and Bradbury et al. we classified our study participants into six different dietary groups (high meat eaters, low meat eaters, poultry eaters, pescatarians, vegetarians, and vegans) based on their responses to a touchscreen FFQ administered at baseline.^14,15^ From 0 (never) to 5 (once or more per day) the questionnaire assessed the consumption of 29 food items—such as various types of meat (e.g., processed meat, lamb, pork, beef, chicken, turkey), fish (oily and non-oily), eggs, and cheese.

When reporting to eat red or processed meat (including poultry) more than 5–6 times per week participants were classified as high meat eaters. Low meat eaters consumed these items less frequently, but more than once a week (Supplementary Table S1). Poultry eaters reported eating poultry but not red or processed meat. Pescatarians excluded red, processed meat, and poultry but included fish in their diet. Vegetarians consumed no meat or fish, while vegans reported never consuming any animal-derived foods. ^14,15^ However, after the initial classification, we found that the number of participants adhering to a vegan diet was too small to allow for statistically robust analyses. To address this limitation, we merged, as previously done by Tong et al. vegetarians and vegans into a single group, referred to as vegetarians ^16^. Additionally, data on average nutrient intakes (e.g., source of protein, fiber, potassium etc.) was derived from the validated Oxford WebQ 24-hour dietary recall questionnaire. ^17^

### Case Ascertainment

CKD diagnoses were identified using the International Classification of Diseases, 10th Revision (ICD-10) codes (N03, N06, N08, N11–N16, N18, N19, Z49, I12, I13) or the Office of Population Censuses and Surveys Classification of Interventions and Procedures, version 4 (OPCS-4), from hospital inpatient data (L74.1–L74.6, L74.8, L74.9, M01.2, M01.3–M01.5, M01.8, M01.9, M02.3, M08.4, M17.2, M17.4, M17.8, M17.9, X40.1–X40.9, X41.1, X41.2, X41.8, X41.9, X42.1, X42.8, X42.9, and X43.1).Full details can be found in Supplementary Table S2.

We classified CKD as prevalent diagnosis if (1) a diagnosis occurred prior to the initial dietary assessment, based on self-reported information and hospital inpatient records, or (2) participants had a baseline estimated glomerular filtration rate (eGFR) below 60 mL/min/1.73 m².

### Covariate Assessment

The following list of covariates was assessed and obtained at baseline recruitment: sex (categorical: Male; Female), age (continuous: Years), type 2 diabetes diagnosis (categorical: Yes; No), ethnicity (categorical: Asian, Black, Mixed, White, Unknown), income before tax (categorical: Less than 18,000, 18,000 to 30,999, 31,000 to 51,999, 52,000 to 100,000, Greater than 100,000, Unknown), education (categorical: High, Medium, Low, Unknown), smoking status (categorical: Never, Previous, Current), alcohol frequency (categorical: Prefer not to answer, Daily or almost daily, Three or four times a week, once or twice a week, One to three times a month, Special occasions only, Never), Body Mass Index (BMI) (continuous: kg/m^2^), waist circumference (continuous: mm), physical activity (categorical: High, Moderate, Low, Unknown), albumin-creatinine ratio (ACR) (categorical: >300 mg/g, 300-30 mg/g, <30 mg/g), eGFR (continuous: ml/min per 1.73m^2^), polygenic risk (High, Medium, Low). The eGFR was calculated using the R package kidney.epi, which utilizes the creatinine-based CKD-EPI (Chronic Kidney Disease Epidemiology Collaboration) 2021 equation. ^18^ The ACR was calculated as follows. ^19^

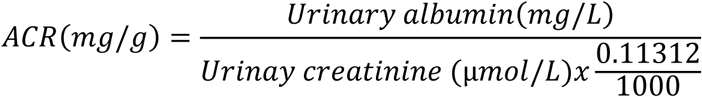

As proposed by the Kidney Disease Improving Global Outcomes (KDIGO) we divided CKD into 5 stages (Supplementary Table S3). The polygenic risk for eGFR was adapted as proposed by Thompson et al. whereby a high polygenic risk score for eGFR reflects a low polygenic risk for CKD. ^20^ Accordingly, we classified participants into three groups: high (high risk for CKD with a low polygenic risk score eGFR), medium (intermediate risk for CKD with a low polygenic eGFR risk score), and low (low risk for CKD with a high polygenic eGFR risk score). A table of all covariates and their respective coding and item number from the UK Biobank’s online showcase can be found in Supplementary Table S4.

### Statistical Analysis

Descriptive statistics were examined across five dietary groups (high meat eaters, low meat eaters, poultry eaters, pescatarians, vegetarians). Continuous variables were expressed as means with standard deviations (±SD), while categorical variables were summarized as percentages. Associations between dietary patterns and incident of CKD were assessed using Cox proportional hazards regression models, with age as the underlying time scale, implemented via the R packages survival and survminer. ^21,22^

Covariates were selected based on a comprehensive review of the literature on established and potential risk factors for CKD, informed by studies such as Charles et al.^23^, Cheung et al.^24^ and Wang et al. ^25^. Primary analyses were adjusted for sociodemographic factors (sex, ethnicity, income after tax, education), lifestyle factors (smoking status, alcohol frequency, physical activity), and clinical measures (T2DM, BMI, waist circumference). Subgroup analyses were conducted stratified by (i) kidney function at baseline (normal vs. CKD stage 1-2), (ii) sex (female vs. male), (iii) polygenic risk (low vs. medium vs. high), (iv) BMI (< 25 vs. ≥ 25), and (v) education (low, medium, high). In sensitivity analyses, models were further adjusted for (i) ACR and eGFR, and (ii) a polygenic risk.

Hazard ratios (HRs) and 95% confidence intervals (CIs) were estimated using Cox proportional hazards models. The proportional hazards assumption was assessed with Schoenfeld residuals and was not violated for the main exposures. Since the assumption was not met for ethnicity and T2DM, these variables were stratified in the models. To evaluate heterogeneity across HRs, Cochrane’s Q and corresponding p-values were calculated using the R package metafor.^26^ A two-sided p-value of less than 0.05 was considered indicative of statistical significance. All analyses were conducted using R (version 4.3.1).^27^

## Results

### 1. Study population

Over a mean (±SD) follow-up period of 12.9 (±2.8) years, 23,084 participants developed CKD among them 90% had normal kidney function at baseline, while 3% and 7% had CKD stage 1 and stage 2, respectively (Supplementary Table S4). Individuals who developed CKD had a higher m ean BMI compared to those who did not (29.0 ± 5.2 vs. 27.2 ± 4.7). Moreover, 27% of participants with type 2 diabetes developed CKD, compared to 7% of those without a history of type 2 diabetes (Supplementary Table S5). Within our study population, 195,030 participants were high mea t eaters, 199,478 low meat eaters, 4,779 poultry eaters, 9,647 pescatarians, and 7,650 vegetarians. When comparing dietary groups, individuals following a vegetarian diet were more likely to be female, younger, have a lower BMI, a higher baseline eGFR, and a higher educational attainment compared to those adhering to high meat eaters (Table 1).

**Table 1:**
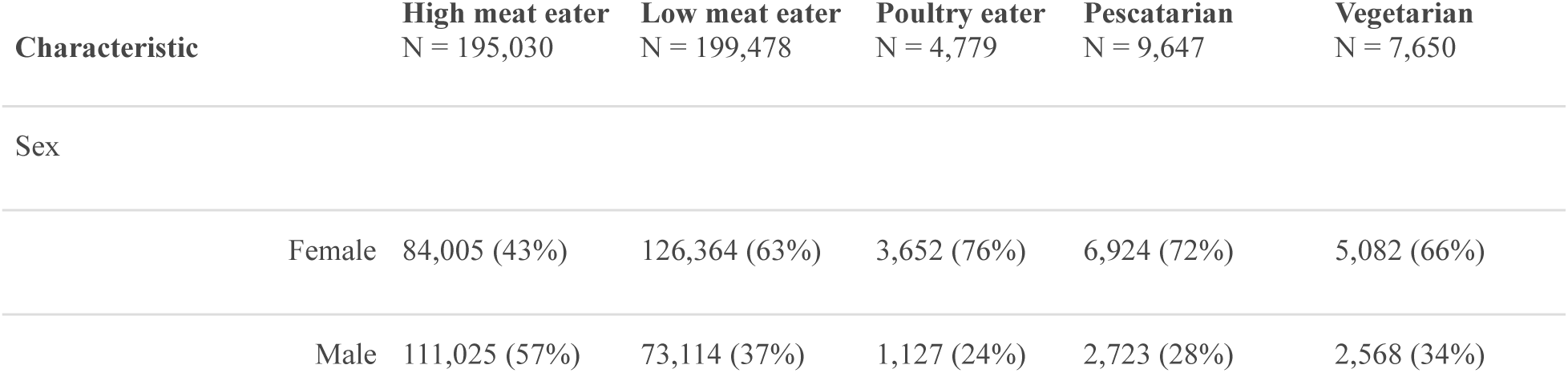
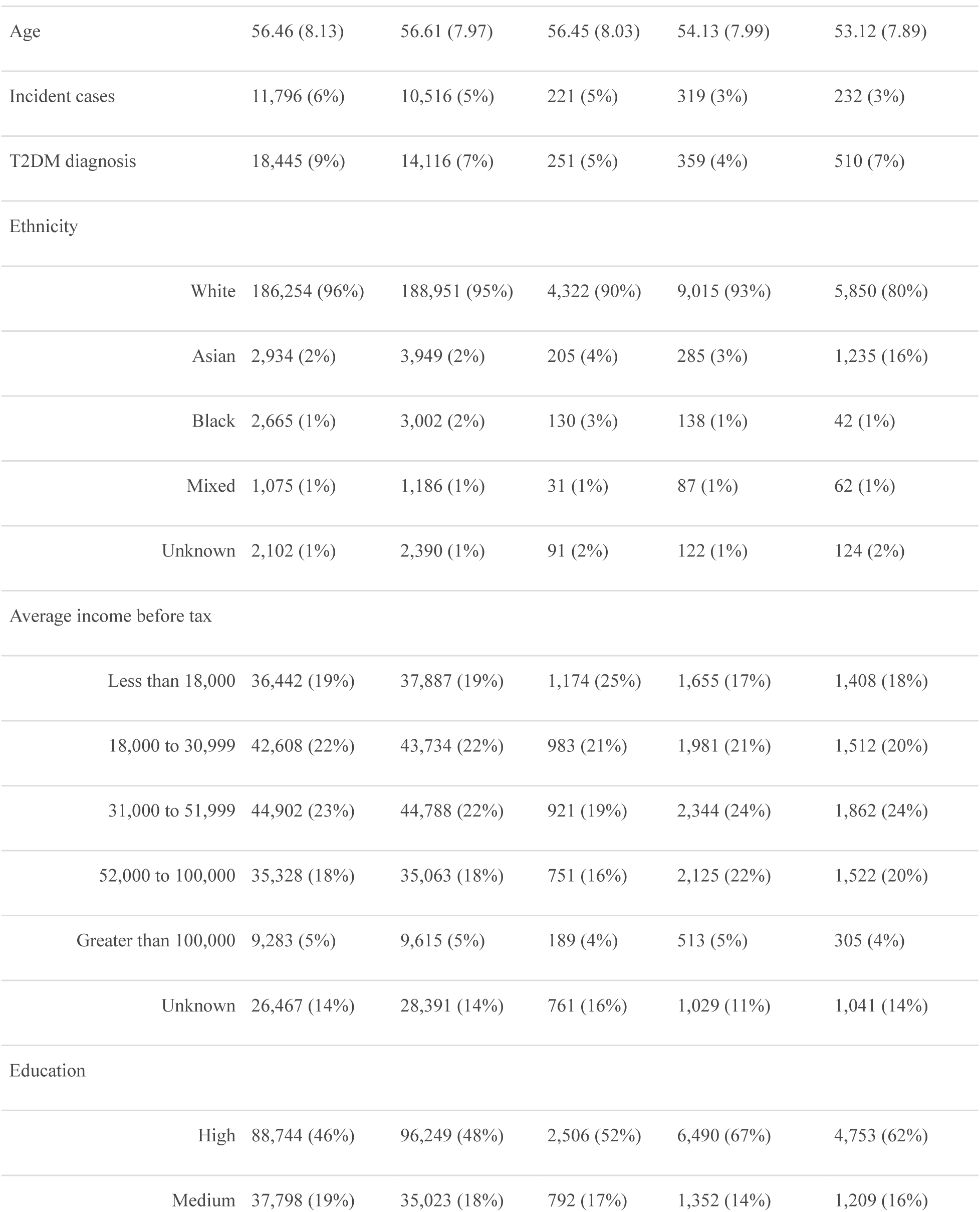
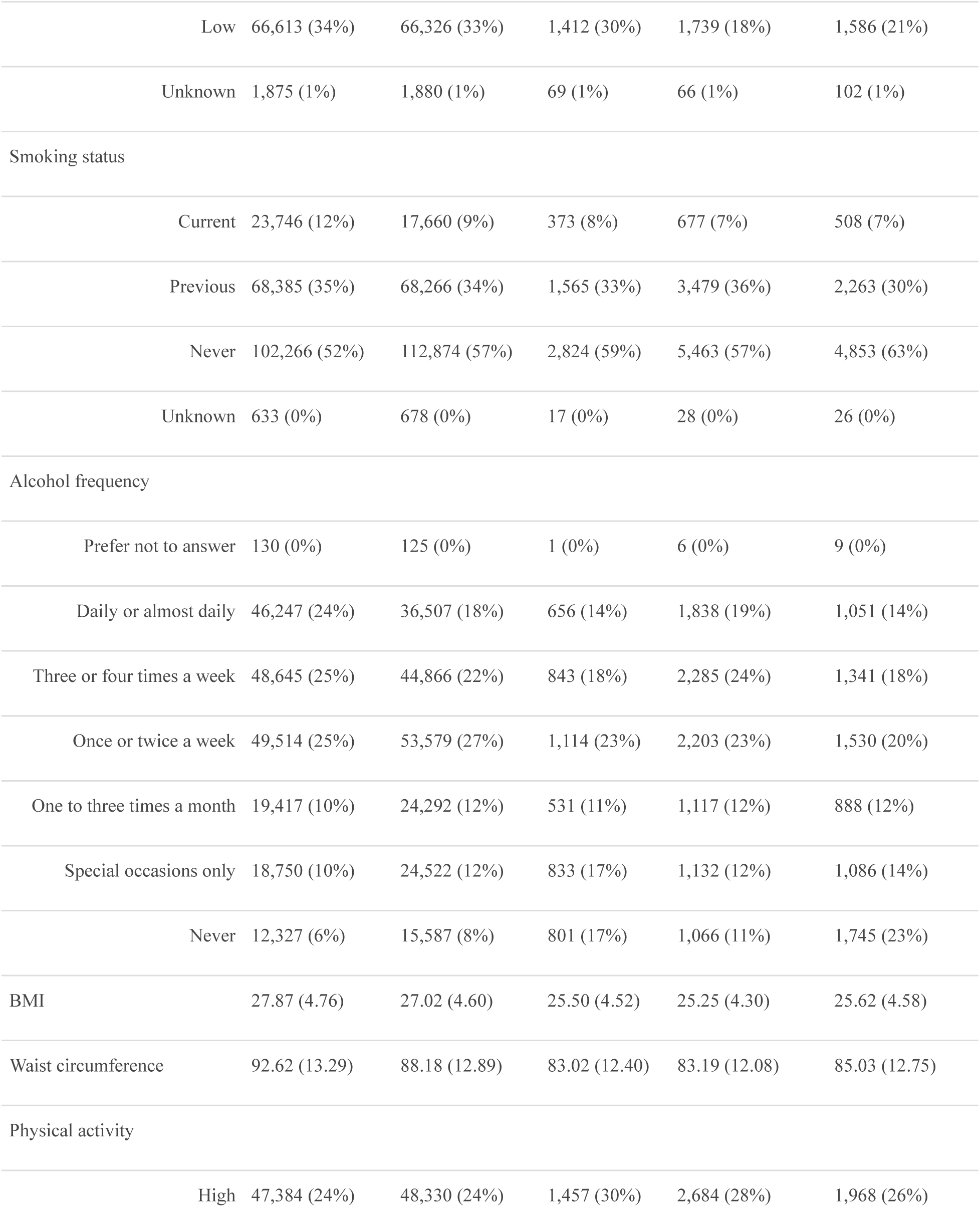
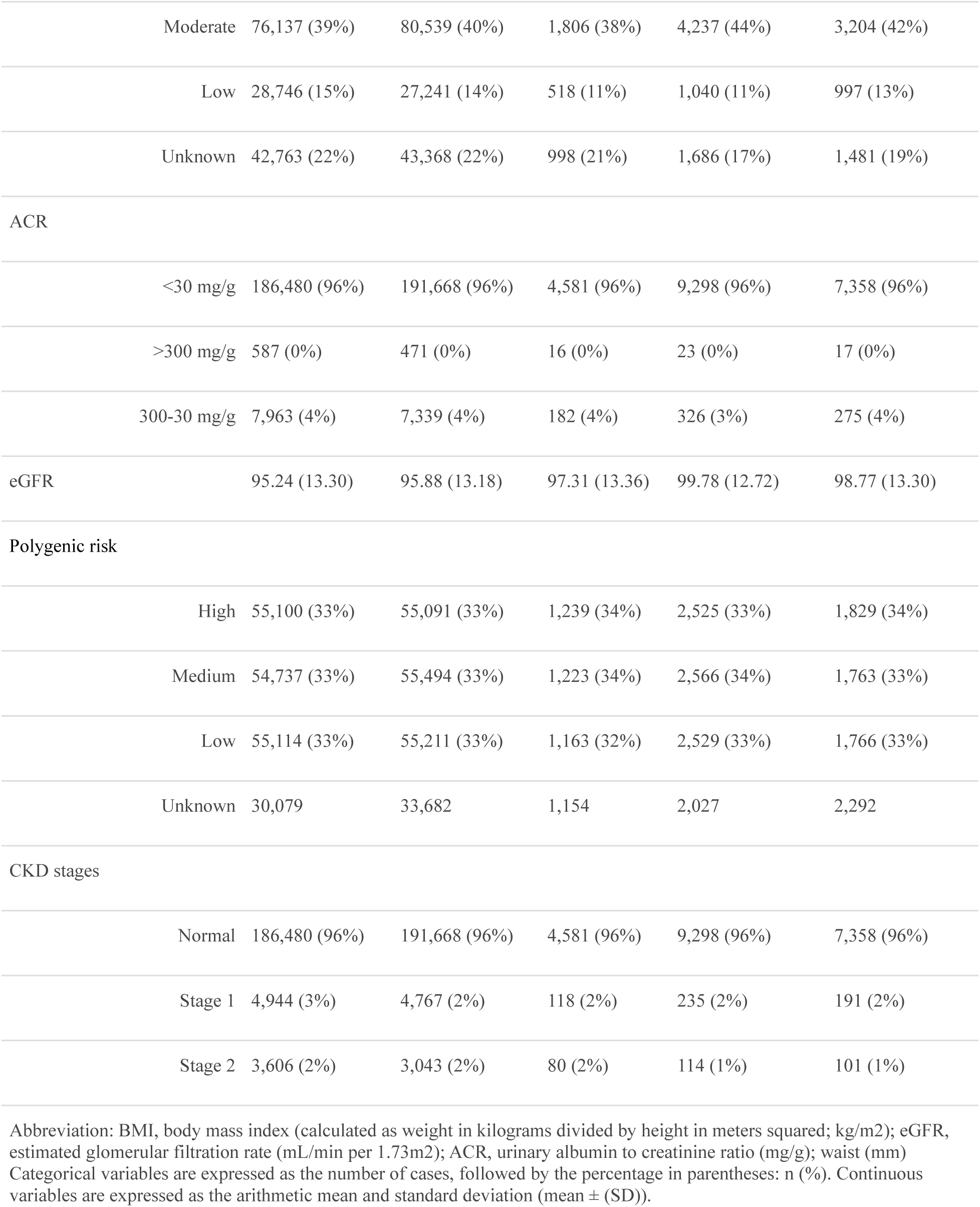
Descriptive statistics among diet groups.

| Characteristic | High meat eater<br>N = 195,030 | Low meat eater<br>N = 199,478 | Poultry eater<br>N = 4,779 | Pescatarian<br>N = 9,647 | Vegetarian<br>N = 7,650 |
| --- | --- | --- | --- | --- | --- |
| Sex |  |  |  |  |  |
| Female | 84,005 (43%) | 126,364 (63%) | 3,652 (76%) | 6,924 (72%) | 5,082 (66%) |
| Male | 111,025 (57%) | 73,114 (37%) | 1,127 (24%) | 2,723 (28%) | 2,568 (34%) |

| Characteristic |  | High meat eater<br>N = 195,030 | Low meat eater<br>N = 199,478 | Poultry eater<br>N = 4,779 | Pescatarian<br>N = 9,647 | Vegetarian<br>N = 7,650 |
| --- | --- | --- | --- | --- | --- | --- |
| Age |  | 56.46 (8.13) | 56.61 (7.97) | 56.45 (8.03) | 54.13 (7.99) | 53.12 (7.89) |
| Incident cases |  | 11,796 (6%) | 10,516 (5%) | 221 (5%) | 319 (3%) | 232 (3%) |
| T2DM diagnosis |  | 18,445 (9%) | 14,116 (7%) | 251 (5%) | 359 (4%) | 510 (7%) |
| Ethnicity |  |  |  |  |  |  |
|  | White | 186,254 (96%) | 188,951 (95%) | 4,322 (90%) | 9,015 (93%) | 5,850 (80%) |
|  | Asian | 2,934 (2%) | 3,949 (2%) | 205 (4%) | 285 (3%) | 1,235 (16%) |
|  | Black | 2,665 (1%) | 3,002 (2%) | 130 (3%) | 138 (1%) | 42 (1%) |
|  | Mixed | 1,075 (1%) | 1,186 (1%) | 31 (1%) | 87 (1%) | 62 (1%) |
|  | Unknown | 2,102 (1%) | 2,390 (1%) | 91 (2%) | 122 (1%) | 124 (2%) |
| Average income before tax |  |  |  |  |  |  |
|  | Less than 18,000 | 36,442 (19%) | 37,887 (19%) | 1,174 (25%) | 1,655 (17%) | 1,408 (18%) |
|  | 18,000 to 30,999 | 42,608 (22%) | 43,734 (22%) | 983 (21%) | 1,981 (21%) | 1,512 (20%) |
|  | 31,000 to 51,999 | 44,902 (23%) | 44,788 (22%) | 921 (19%) | 2,344 (24%) | 1,862 (24%) |
|  | 52,000 to 100,000 | 35,328 (18%) | 35,063 (18%) | 751 (16%) | 2,125 (22%) | 1,522 (20%) |
|  | Greater than 100,000 | 9,283 (5%) | 9,615 (5%) | 189 (4%) | 513 (5%) | 305 (4%) |
|  | Unknown | 26,467 (14%) | 28,391 (14%) | 761 (16%) | 1,029 (11%) | 1,041 (14%) |
| Education |  |  |  |  |  |  |
|  | High | 88,744 (46%) | 96,249 (48%) | 2,506 (52%) | 6,490 (67%) | 4,753 (62%) |
|  | Medium | 37,798 (19%) | 35,023 (18%) | 792 (17%) | 1,352 (14%) | 1,209 (16%) |
|  | Low | 66,613 (34%) | 66,326 (33%) | 1,412 (30%) | 1,739 (18%) | 1,586 (21%) |
|  | Unknown | 1,875 (1%) | 1,880 (1%) | 69 (1%) | 66 (1%) | 102 (1%) |
| Smoking status |  |  |  |  |  |  |
|  | Current | 23,746 (12%) | 17,660 (9%) | 373 (8%) | 677 (7%) | 508 (7%) |
|  | Previous | 68,385 (35%) | 68,266 (34%) | 1,565 (33%) | 3,479 (36%) | 2,263 (30%) |
|  | Never | 102,266 (52%) | 112,874 (57%) | 2,824 (59%) | 5,463 (57%) | 4,853 (63%) |
|  | Unknown | 633 (0%) | 678 (0%) | 17 (0%) | 28 (0%) | 26 (0%) |
| Alcohol frequency |  |  |  |  |  |  |
|  | Prefer not to answer | 130 (0%) | 125 (0%) | 1 (0%) | 6 (0%) | 9 (0%) |
|  | Daily or almost daily | 46,247 (24%) | 36,507 (18%) | 656 (14%) | 1,838 (19%) | 1,051 (14%) |
|  | Three or four times a week | 48,645 (25%) | 44,866 (22%) | 843 (18%) | 2,285 (24%) | 1,341 (18%) |
|  | Once or twice a week | 49,514 (25%) | 53,579 (27%) | 1,114 (23%) | 2,203 (23%) | 1,530 (20%) |
|  | One to three times a month | 19,417 (10%) | 24,292 (12%) | 531 (11%) | 1,117 (12%) | 888 (12%) |
|  | Special occasions only | 18,750 (10%) | 24,522 (12%) | 833 (17%) | 1,132 (12%) | 1,086 (14%) |
|  | Never | 12,327 (6%) | 15,587 (8%) | 801 (17%) | 1,066 (11%) | 1,745 (23%) |
| BMI |  | 27.87 (4.76) | 27.02 (4.60) | 25.50 (4.52) | 25.25 (4.30) | 25.62 (4.58) |
| Waist circumference |  | 92.62 (13.29) | 88.18 (12.89) | 83.02 (12.40) | 83.19 (12.08) | 85.03 (12.75) |
| Physical activity |  |  |  |  |  |  |
|  | High | 47,384 (24%) | 48,330 (24%) | 1,457 (30%) | 2,684 (28%) | 1,968 (26%) |
|  | Moderate | 76,137 (39%) | 80,539 (40%) | 1,806 (38%) | 4,237 (44%) | 3,204 (42%) |
|  | Low | 28,746 (15%) | 27,241 (14%) | 518 (11%) | 1,040 (11%) | 997 (13%) |
|  | Unknown | 42,763 (22%) | 43,368 (22%) | 998 (21%) | 1,686 (17%) | 1,481 (19%) |
| ACR |  |  |  |  |  |  |
|  | <30 mg/g | 186,480 (96%) | 191,668 (96%) | 4,581 (96%) | 9,298 (96%) | 7,358 (96%) |
|  | >300 mg/g | 587 (0%) | 471 (0%) | 16 (0%) | 23 (0%) | 17 (0%) |
|  | 300-30 mg/g | 7,963 (4%) | 7,339 (4%) | 182 (4%) | 326 (3%) | 275 (4%) |
| eGFR |  | 95.24 (13.30) | 95.88 (13.18) | 97.31 (13.36) | 99.78 (12.72) | 98.77 (13.30) |
| Polygenic risk |  |  |  |  |  |  |
|  | High | 55,100 (33%) | 55,091 (33%) | 1,239 (34%) | 2,525 (33%) | 1,829 (34%) |
|  | Medium | 54,737 (33%) | 55,494 (33%) | 1,223 (34%) | 2,566 (34%) | 1,763 (33%) |
|  | Low | 55,114 (33%) | 55,211 (33%) | 1,163 (32%) | 2,529 (33%) | 1,766 (33%) |
|  | Unknown | 30,079 | 33,682 | 1,154 | 2,027 | 2,292 |
| CKD stages |  |  |  |  |  |  |
|  | Normal | 186,480 (96%) | 191,668 (96%) | 4,581 (96%) | 9,298 (96%) | 7,358 (96%) |
|  | Stage 1 | 4,944 (3%) | 4,767 (2%) | 118 (2%) | 235 (2%) | 191 (2%) |
|  | Stage 2 | 3,606 (2%) | 3,043 (2%) | 80 (2%) | 114 (1%) | 101 (1%) |

Abbreviation: BMI, body mass index (calculated as weight in kilograms divided by height in meters squared; kg/m<sup>2</sup>); eGFR, estimated glomerular filtration rate (mL/min per 1.73m<sup>2</sup>); ACR, urinary albumin to creatinine ratio (mg/g); waist (mm). Categorical variables are expressed as the number of cases, followed by the percentage in parentheses: n (%). Continuous variables are expressed as the arithmetic mean and standard deviation (mean ± (SD)).

Nutrient intake varied across dietary groups. High meat consumers had the highest intake of ani mal-based protein, whereas vegetarians had the highest intake of plant-based protein. Vegetarian s also consumed more fiber but less potassium and phosphate compared to high meat eaters (Supplementary Table S6). Overall, meat eaters had higher urinary excretion of sodium, creatinine, an d microalbumin than non–meat eaters (Supplementary Table S7).

### 2. Diet group and risk of CKD

In fully adjusted models (Figure 1), low meat eaters [HR = 0.97, 95% CI: 0.94– 1.00] and vegetarians [HR = 0.81, 95% CI: 0.71–0.93] had a significantly lower risk of CKD compared with high meat eaters.

**Figure 1:**
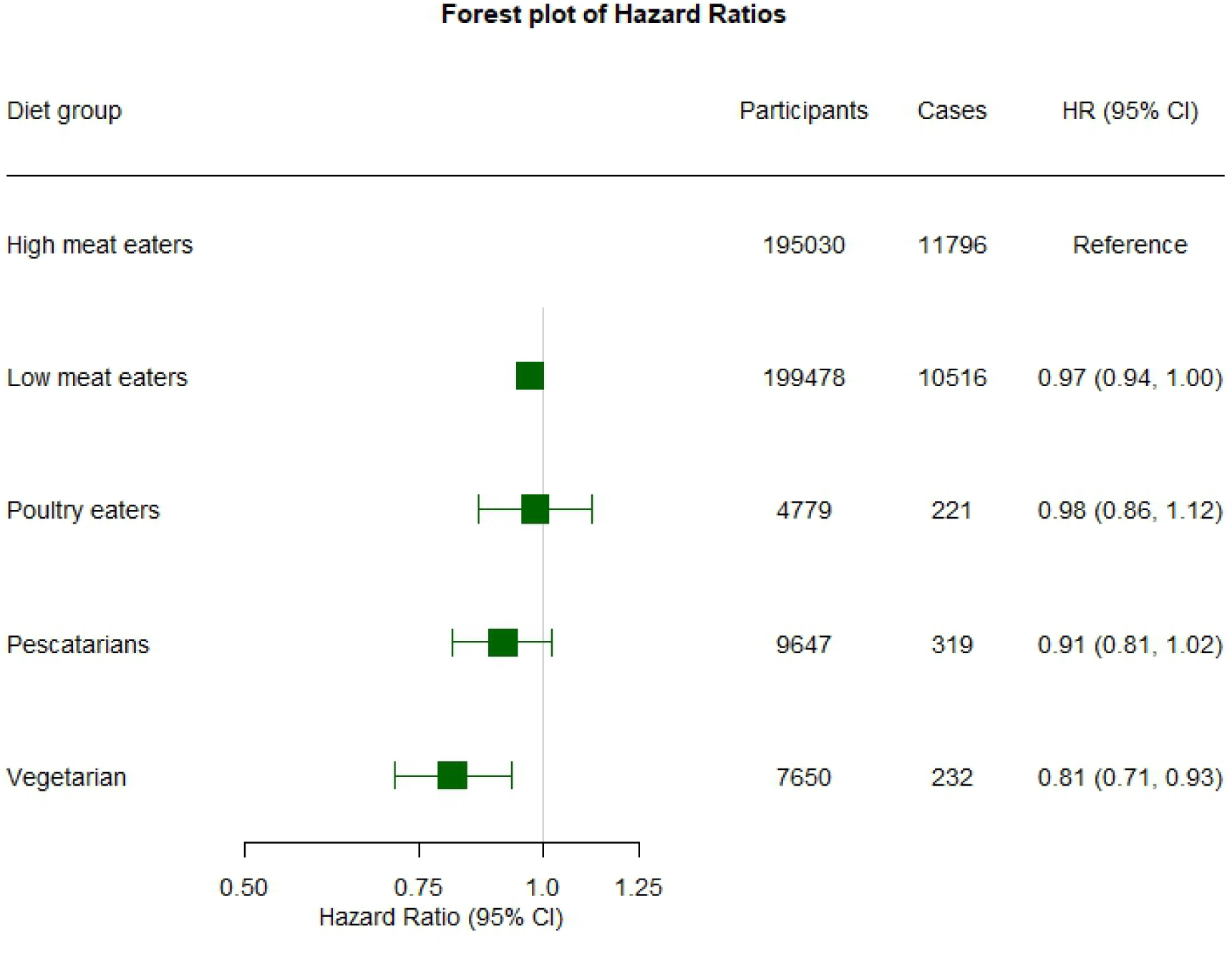
Hazard ratios (HR) and 95% Confidence intervals (95% CI) between diet groups and the risk of chronic kidney disease (CKD). Age included as the underlying time variable. Models adjusted for sex, income and education level, waist circumference, body mass index (BMI), physical activity and smoking status and stratified by ethnicity and type 2 diabetes mellitus (T2DM) diagnosis.

Stratification by baseline kidney function showed that vegetarians with normal kidney function had a lower risk of CKD [HR = 0.81, 95% CI: 0.70–0.93], while no significant association was observed among those with mildly reduced kidney function (CKD stages 1-2); however, there was no evidence of heterogeneity (p=0.27-0.84, Supplementary Figure S2). The inverse association for vegetarian diets was slightly more pronounced in men (21% lower risk, 95% CI: 0.64 – 0.98) than in women (16% lower risk, 95% CI: 0.71 – 0.99), but no significant heterogeneity by sex was observed (Supplementary Figure S3). Participants within the lowest polygenic risk group (indicative of higher kidney function) appeared to benefit most, with a ∼30% lower risk of CKD compared with high meat eaters, although the associations were consistent across polygenic risk strata and no heterogeneity was detected (p=0.35 to 0.95) (Supplementary Figure S4). In subgroup analyses across different BMI and educational groups, varying trends were observed. However, tests for heterogeneity did not reach statistical significance (Supplementary Figure S5-S6).

In sensitivity analyses with additional adjustment for baseline eGFR and ACR, the association observed among vegetarians was slightly strengthened [HR = 0.80, 95% CI: 0.68–0.94] (Supplementary Figure S7). After further adjustment for polygenic risk, the association for pescatarians became stronger and reached statistical significance [HR = 0.89, 95% CI: 0.75–0.97], while the association for vegetarians remained similar [HR = 0.79, 95% CI: 0.67–0.94] (Supplementary Figure S8).

## Discussion

In this large population-based study, we observed a gradient in CKD risk reduction across low meat eaters (3% lower), poultry eaters (2% lower), pescatarians (9%) and vegetarians (19% lower) compared to high meat eaters. These associations paralleled a gradient in animal protein intake and were consistent across population subgroups, suggesting that vegetarian diets may provide the greatest protection against CKD.

While diets rich in fruits and vegetables confer health benefits in the general population^8,28,29^, recommendations for individuals with CKD can be contradictory. Protein restriction, used to slow CKD progression, favors diets low in animal products, whereas potassium-restricted diets to prevent hyperkalemia limit many plant-based foods. Additionally, dietary phosphate restriction—key to managing CKD-related mineral and bone disorders —often leads to monotonous diets with limited variety.

Our results are in line with findings from a US cohort study, which categorised 14,686 participants into overall, healthy, and less healthy plant-based diet indices, as well as a pro-vegetarian diet index. Over a median follow-up of 24 years, almost 30% developed incident CKD, with risk 10-14 % lower among those adhering to a pro-vegetarian diet and healthy plant-based diets. Of note, CKD incidence was considerably higher in *this population* than in the UK Biobank, likely reflecting differences in follow-up length, cohort composition, and baseline risk^9^. Similarly, in a Taiwanese cohort with self-reported diet groups, individuals following plant-based diets had lower CKD risk.^30^ Prior studies using dietary pattern and diet quality scores have consistently shown that greater inclusion of plant-based foods is associated with lower incidence of CKD^31^, slower disease progression, and reduced mortality among CKD patients.^6,32,33^ Interestingly, we show that, although the strength of the association was slightly weaker in those with a lower genetic risk for CKD, there was no heterogeneity across strata of genetic risk, indicating regardless of genetic risk such dietary patterns may be beneficial.

The biological mechanisms through which plant-based diets might mitigate the development of kidney disease are not elucidated in detail. However, three main mechanisms might positively influence the development and progression of CKD, respectively.

(1) Plant-based diets are usually low in organic phosphate compounds, organic sulfate, and non-metabolizable organic acids, which lead to proton excess and subsequent acid loading.^34,35^ Experimental studies have shown that renal tissue acidification enhances the local renin-angiotensin system and that angiotensin II, aldosterone and endothelin promote renal fibrosis, leading to progression of CKD.^35,36^ Clinical studies in patients with hypertensive nephropathy showed that a diet rich in fruits and vegetables reduced net acid excretion and halted the progression of CKD, when compared to drug-based intervention (and add in the drugs used).^37–39^
(2) Plant-based diets are come with a higher dietary intake of fiber and polyphenols. Polyphenols have previously been associated with a lower risk of CKD and a reduced all-cause mortality in patients with CKD.^40,41^ Higher dietary fiber intake supports a healthier gut microbiome and is associated with a lower incidence of CKD^42–44^ and mortality,^45^ possibly due to an increase in short-chain fatty acid (SCFA)-producing bacteria, which have been shown to reduce oxidative stress and inflammation.^46^ Plant-based diet may also lower circulating trimethylamine-N-oxide (TMAO) levels, a gut-derived metabolite associated with higher mortality and cardiometabolic risk in CKD ^47^ – thus, TMAO may be a target for therapeutic strategies in CKD. ^48–50^ In addition, comorbidities strongly associated with CKD, such as diabetes or hypertension, are less common with higher fiber intake.^51^ Recently, it has been shown that dietary fiber intake reduces the development of diabetic nephropathy *via* an increased number of SCFA-producing bacteria,^52^ and halts the progression of CKD-MBD in mouse models.^53^ The gut-kidney axis appears to play a pivotal role in the development of CKD and associated comorbidities,^54^ and thus diet represents a modifiable risk factor to prevent these diseases and mitigate their progression.
(3) Micronutrients are present in reasonable concentrations in plant-based meals. While effects of minerals such as potassium and phosphate on kidney function have been analysed in detail, the effects of other micronutrients are less well understood. In the *Tehran Lipid and Glucose Study (TLGS* ), the risk of incident CKD was lower with higher intakes of vitamins C, E, D, cobalamin, and folate.^55^ This partly contrasts with the findings of the *Ansan-Ansung cohort* of *the Korean Genome and Epidemiologic Study (KoGES*), in which participants with higher vitamin C and B6 intakes had a higher risk of developing CKD over a 12-year follow-up period.^56^ However, beneficial micronutrient intake may reduce oxidative stress and inflammation and subsequently modify the progression of CKD.^57^ Interestingly, high potassium intake was associated with a lower incidence in CKD in a recent *UK Biobank* analysis.^58^ Compared with individuals in the lowest quintile of dietary potassium intake, those in the highest quintile had a 14% lower risk of incident CKD. This association was even more pronounced when urinary potassium-to-creatinine excretion was used as a surrogate marker of potassium intake. High quality plant-based diets are high in potassium and low in sodium. The detrimental effects of high sodium intake are well known, and the effect of a low-sodium diet may also partly explain our findings.^59,60^ Similarly, red meat–rich diets are high in phosphate, a key driver of CKD–mineral and bone disorder (CKD-MBD), which in turn is a major contributor to the excess mortality observed in CKD populations.^9^ In addition, higher dietary phosphate intake is also associated with higher mortality in the general population.^61^ In plant foods, phosphate occurs mainly as phytate, which is poorly absorbed due to the lack of intestinal phytase, thereby reducing phosphate burden.^62–64^

This study has several limitations. The UK Biobank cohort is characterized by a generally healthier risk profile relative to the UK general population and predominantly composed of participants of white European descent.^65^ This may limit the generalizability of the findings to more diverse populations. Another key limitation relates to its observational design, which precludes the exclusion of residual or unmeasured confounding. Although a considerable number of CKD cases were recorded during follow-up, the statistical power to detect moderate associations among vegans was limited due to the relatively small size of this subgroup (n=367). However, vegan and vegetarian diets have partially distinct nutritional profiles, which may lead to a dilution of associations when combining them. ^66^ Lastly, participants may have changed their dietary habits over time or underreported certain food items, which may lead to dietary misclassification, which in fact may have led to an underestimation of the true risk estimates. However, a reproducibility study in a subsample of the UK Biobank suggests that diet types remain very stable over time in this cohort.^67^

To the best of our knowledge this is the first study that shows, that vegetarian diets are associated with a lower risk of CKD. This finding is consistent with previous research highlighting the benefits of plant-based dietary patterns, as captured by various dietary indices ^6,7,32^. Notably, recent clinical guidelines such as those from KDIGO (Kidney Disease: Improving Global Outcomes) have begun to acknowledge and support the role of plant-based diets in CKD management ^68^. However, in clinical practice, more conservative approaches—often based on outdated concerns regarding potassium, phosphorus, or protein quality—still prevail.^69^ Given the potential benefit of plant-based diets in the prevention of CKD, randomized controlled trials are needed on plant-based diets and prognosis among CKD patients, including assessing their practicability and acceptability.

## Data Availability

This research was conducted using data from UK Biobank under application number 64226. The data are not publicly available but can be accessed by bona fide researchers upon application to UK Biobank.

## Acknowledgements

This research has been conducted using the UK Biobank. We are deeply grateful to all the participants who generously contributed their time and data, without whom this study would not have been possible.

## Authors’ Contributions

Research idea and study design: CJC, MG, TK; data acquisition: WB, AST; analysis/interpretation: CJC, MG, TK, AC, SR; statistical analysis: CJC, WB, AST; supervision or mentorship: TK, MG, AC, SR. Each author contributed important intellectual content during manuscript drafting or revision and agrees to be personally accountable for the individual’s own contributions and to ensure that questions pertaining to the accuracy or integrity of any portion of the work, even one in which the author was not directly involved, are appropriately investigated and resolved, including with documentation in the literature if appropriate.

## Data Sharing Statement

This research was conducted using data from UK Biobank under application number 64226. The data are not publicly available but can be accessed by bona fide researchers upon application to UK Biobank (www.ukbiobank.ac.uk).

## Ethics approval and consent to participate

Ethical approval for the UK Biobank cohort was obtained from the National Health Service North West Multi-Centre Research Ethics Committee (21/NW/0157). Written informed consent was received from all participants. Further details regarding the study protocol and data access for researchers have been published elsewhere (https://www.ukbiobank.ac.uk/media/gnkeyh2q/study-rationale.pdf).

## Declaration of generative AI in scientific writing

During the preparation of this work the author(s) used DeepL in order to improve the clarity and readability of the text. After using this tool/service, the author(s) reviewed and edited the content as needed and take(s) full responsibility for the content of the published article.

