## Supplementary for "Incident Chronic Kidney Disease in meat-eaters, fish-eaters, and vegetarians: A population-based prospective study"

***Supplementary Figure 1:*** *Flow chart of included participants*

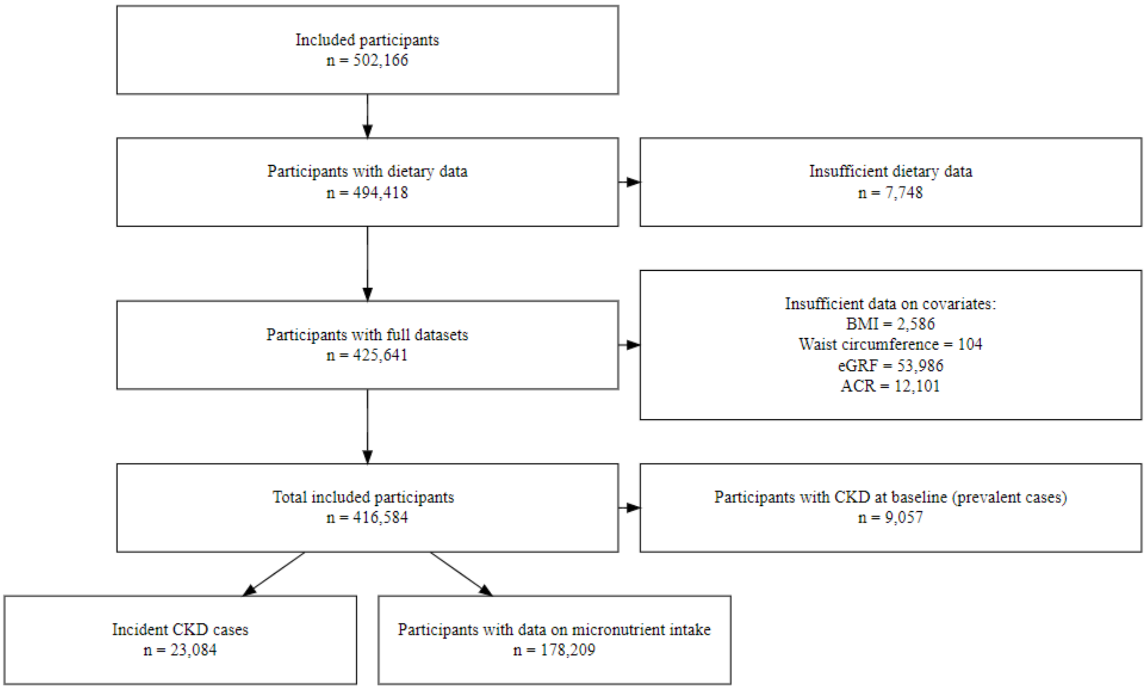

***Supplementary Table 1****: Diet classification*

| | High meat eater | Low meat eater | Poultry eater | Pescetarian | Vegetarian |
|---|---|---|---|---|---|
| Processed meat | | | | | |
| Beef | | | | | |
| Pork | More than 5 times per week | 1 – 4 times per week | | | |
| Lamp, mutton | | | | | |
| Chicken, turkey, poultry | | | | | |
| Fish | | | | | |
| Egg | | | | | |

| |
|---|
| Dairy |

***Supplementary Table 2:*** *Cases assessment of chronic kidney disease (CKD)*

| Code Type | Code | UK Biobank Showcase Data Field |
|---|---|---|
| Self-reported non-cancer illness | 1192, 1193, 1194, 1607 | 20002 |
| ICD-9 codes | 585.9 | 41271 |
| ICD-10 codes | N03, N06, N08, N11, N12, N13, N14, N15, N16, N18, N19, Z49, I12, I13 | 41270 |
| OPCS-3 codes | 560.2, 561, 561.1, 562, 563.1, 563.2, 563.3, 564, 564.1, 565, 566, 566.1, 567.1, 567.2, 568, 569, 571.1 | 41273 |
| OPCS-4 codes | L74.1, L74.2, L74.3, L74.4, L74.5, L74.6, L74.8, L74.9, M01.2, M01.3, M01.4, M01.5, M01.8, M01.9, M02.3, M08.4, M17.2, M17.4, M17.8, M17.9, X40.1, X40.2, X40.3, X40.4, X40.5, X40.6, X40.7, X40.8, X40.9, X41.1, X41.2, X41.8, X41.9, X42.1, X42.8, X42.9, X43.1 | 41272 |

***Supplementary Table 3:*** *CKD Stage classification adapted from KDIGO (Kidney Disease: Improving Global Outcomes )*[1]

| CKD Stage | eGFR | ACR |
|---|---|---|
| Normal kidney function | > 90 | <= 30 |
| Stage 1 | > 90 | > 30 |
| Stage 2 | >= 60 & <= 90 | > 30 |
| Stage 3 | >= 30 & < 60 | |
| Stage 4 | >= 15 & < 30 | |
| Stage 5 | < 15 | |

Abbreviation: eGFR, estimated glomerular filtration rate (mL/min per 1.73m2); ACR, urinary albumin to creatinine ratio (mg/g);

***Supplementary Table 4:*** *Information on categorization of covariates*

| Variables | UK Biobank Showcase Data Field | Type | Coding |
|---|---|---|---|
| Sex | 31 | Categorical: male, female | NA |
| Age | 21022 | Continuous: years | NA |
| T2DM Diagnosis | 1712 | Categorical: Yes, No | ICD10 E11 Codes |
| Ethnicity | 100065 | Categorical: Asian, Black, Mixed, White, Unknown | NA |
| Income before tax | 738 | Categorical: „Less than 18,000“, „18,000 to 30,999“ , | NA |

| | | | |
|---|---|---|---|
| | | „31,000 to 51,999“, “52,000 to 100,000”, “Greater than 100,000”, “Missing” | |
| Education | 6138 | Categorical: Low, Medium, High, Unknown | Low = “None of the above”, “O-level/GCSE”, “cse or equivalent”<br><br>Medium = “A-level/AS Level”, “NVQ, HND, HNC”<br><br>High = “College or University Degree”, “Other Professional Qualification”<br><br>Unknown = „prefer not to answer“, missing, „none of the above“ |
| Smoking status | 20116 | Categorical: Never, previous, Current, Unknown | NA |
| Alcohol Frequency | 1558 | categorical: prefer not to answer, daily or almost daily, three or four times a week, once or twice a week, one to three times a month, special occasions only, never | NA |
| BMI (Body Mass Index) | 23104 | Continuous: kg/m2 | NA |
| Waist circumference | 48 | continuous: cm | NA |
| Physical activity | 874, 894, 914 | categorical: high, moderate, low, unknown | Low = under 600 minutes/week<br>Moderate = 600-3000 minutes/week<br>High = over 3000 minutes/week |
| ACR (albumin creatinine ratio) | 30510, 30505 | categorical:<br><30 mg/g,<br>30-300 mg/g,<br>>300 mg/g | |
| eGFR (estimated glomerular filtration rate) | 23478, 21000 | continuous: $mL/min/1.73m^2$ | Calculated using creatinine-based CKD-EPI 2021 from the R package kidney.epi [2] |

| Polygenic risk | | Categorical: high, medium, low | Polygenic risk score was calculated based on Thompson et al. and subsequently divided into tertiles[3] |
|---|---|---|---|

***Supplementary Table 5:*** *Descriptive characteristics among incident CKD cases and non-cases*

| | Characteristic | Non- Cases<br>N = 393,500 | Cases<br>N = 23,084 | Overall<br>N = 416,584 |
|---|---|---|---|---|
| Diet | High meat eater | 183,234 (47%) | 11,796 (51%) | 195,030 (47%) |
| | Low meat eater | 188,962 (48%) | 10,516 (46%) | 199,478 (48%) |
| | Poultry eater | 4,558 (1%) | 221 (1%) | 4,779 (1%) |
| | Pescatarian | 9,328 (2%) | 319 (1%) | 9,647 (2%) |
| | Vegetarian | 7,418 (2%) | 232 (1%) | 7,650 (2%) |
| Sex | Female | 215,029 (55%) | 10,998 (48%) | 226,027 (54%) |
| | Male | 178,471 (45%) | 12,086 (52%) | 190,557 (46%) |
| Age | | 56.14 (8.06) | 61.02 (6.72) | 56.41 (8.07) |
| T2DM diagnosis | | 27,570 (7%) | 6,111 (26%) | 33,681 (8%) |
| Ethnicity | White | 372,875 (95%) | 21,854 (95%) | 394,729 (95%) |
| | Asian | 8,152 (2%) | 456 (2%) | 8,608 (2%) |
| | Black | 5,581 (1%) | 396 (2%) | 5,977 (1%) |
| | Mixed | 2,336 (1%) | 105 (0%) | 2,441 (1%) |
| | Unknown | 4,556 (1%) | 273 (1%) | 4,829 (1%) |
| Average income before tax | Less than 18,000 | 71,678 (18%) | 6,888 (30%) | 78,566 (19%) |
| | 18,000 to 30,999 | 85,396 (22%) | 5,422 (23%) | 90,818 (22%) |
| | 31,000 to 51,999 | 90,872 (23%) | 3,945 (17%) | 94,817 (23%) |
| | 52,000 to 100,000 | 72,525 (18%) | 2,264 (10%) | 74,789 (18%) |
| | Greater than 100,000 | 19,433 (5%) | 472 (2%) | 19,905 (5%) |
| | Unknown | 53,596 (14%) | 4,093 (18%) | 57,689 (14%) |
| Education | High | 190,188 (48%) | 8,554 (37%) | 198,742 (48%) |
| | Medium | 72,226 (18%) | 3,948 (17%) | 76,174 (18%) |
| | Low | 127,386 (32%) | 10,290 (45%) | 137,676 (33%) |
| | Unknown | 3,700 (1%) | 292 (1%) | 3,992 (1%) |
| | Current | 40,338 (10%) | 2,626 (11%) | 42,964 (10%) |

| | Characteristic | Non- Cases<br>N = 393,500 | Cases<br>N = 23,084 | Overall<br>N = 416,584 |
|---|---|---|---|---|
| Smoking status | Previous | 134,371 (34%) | 9,587 (42%) | 143,958 (35%) |
| | Never | 217,519 (55%) | 10,761 (47%) | 228,280 (55%) |
| | Unknown | 1,272 (0%) | 110 (0%) | 1,382 (0%) |
| Alcohol frequency | Prefer not to answer | 253 (0%) | 18 (0%) | 271 (0%) |
| | Daily or almost daily | 81,870 (21%) | 4,429 (19%) | 86,299 (21%) |
| | Three or four times a week | 93,682 (24%) | 4,298 (19%) | 97,980 (24%) |
| | Once or twice a week | 102,306 (26%) | 5,634 (24%) | 107,940 (26%) |
| | One to three times a month | 43,600 (11%) | 2,645 (11%) | 46,245 (11%) |
| | Special occasions only | 42,888 (11%) | 3,435 (15%) | 46,323 (11%) |
| | Never | 28,901 (7%) | 2,625 (11%) | 31,526 (8%) |
| BMI | | 27.23 (4.66) | 29.04 (5.18) | 27.33 (4.71) |
| Waist circumference | | 89.70 (13.21) | 95.53 (13.88) | 90.02 (13.31) |
| Physical activity | High | 96,632 (25%) | 5,191 (22%) | 101,823 (24%) |
| | Moderate | 157,639 (40%) | 8,284 (36%) | 165,923 (40%) |
| | Low | 54,965 (14%) | 3,577 (15%) | 58,542 (14%) |
| | Unknown | 84,264 (21%) | 6,032 (26%) | 90,296 (22%) |
| ACR category | <30 mg/g | 378,701 (96%) | 20,684 (90%) | 399,385 (96%) |
| | >300 mg/g | 770 (0%) | 344 (1%) | 1,114 (0%) |
| | 300-30 mg/g | 14,029 (4%) | 2,056 (9%) | 16,085 (4%) |
| eGFR | | 96.41 (12.90) | 84.33 (14.03) | 95.74 (13.26) |
| Polygenic risk | High | 108,519 (33%) | 7,265 (37%) | 115,784 (33%) |
| | Medium | 109,335 (33%) | 6,448 (33%) | 115,783 (33%) |
| | Low | 109,949 (34%) | 5,834 (30%) | 115,783 (33%) |
| | Unknown | 65,697 | 3,537 | 69,234 |
| CKD stages | Normal | 378,701 (96%) | 20,684 (90%) | 399,385 (96%) |
| | Stage 1 | 9,548 (2%) | 707 (3%) | 10,255 (2%) |

| Characteristic | Non- Cases<br>N = 393,500 | Cases<br>N = 23,084 | Overall<br>N = 416,584 |
|---|---|---|---|
| Stage 2 | 5,251 (1%) | 1,693 (7%) | 6,944 (2%) |

Abbreviation: BMI, body mass index (calculated as weight in kilograms divided by height in meters squared; kg/m2); eGFR, estimated glomerular filtration rate (mL/min per 1.73m2); ACR, urinary albumin to creatinine ratio (mg/g); waist (mm)
Categorical variables are expressed as the number of cases, followed by the percentage in parentheses: n (%).
Continuous variables are expressed as the arithmetic mean and standard deviation (mean ± (SD)).

***Supplementary Table 6:*** *Nutritional intake among different dietary groups*

| **Characteristic** | **High meat eater**<br>**N = 79,948** | **Low meat eater**<br>**N = 86,976** | **Poultry eater**<br>**N = 2,091** | **Pescatarian**<br>**N = 5,215** | **Vegetarian**<br>**N = 3,979** |
|---|---|---|---|---|---|
| Protein average (g/day) | 85 (24) | 78 (22) | 72 (23) | 68 (20) | 62 (19) |
| Animal protein (g/day) | 57 (21) | 51 (19) | 42 (20) | 33 (16) | 23 (13) |
| Plant-based protein (g/day) | 28 (9) | 27 (10) | 30 (12) | 35 (13) | 40 (15) |
| Potassium average (mg/day) | 3.70 (1.01) | 3.60 (1.02) | 3.65 (1.15) | 3.68 (1.04) | 3.62 (1.10) |
| Sodium average (mg/day) | 2.07 (0.78) | 1.82 (0.69) | 1.73 (0.70) | 1.91 (0.70) | 1.93 (0.71) |
| Phosphate average (mg/day) | 1.45 (0.37) | 1.39 (0.36) | 1.37 (0.39) | 1.39 (0.37) | 1.36 (0.39) |
| Fibre avarage (g/day) | 17 (6) | 18 (6) | 20 (8) | 21 (7) | 22 (8) |
| Vitamin E average (mg/day) | 11.0 (4.3) | 10.6 (4.2) | 11.5 (4.7) | 12.4 (4.6) | 12.6 (5.2) |
| Vitamin D average (ug/day) | 3.71 (2.78) | 3.56 (2.90) | 3.92 (3.58) | 3.82 (3.41) | 2.05 (1.57) |
| Vitamin C average (mg/day) | 122 (74) | 130 (77) | 146 (88) | 143 (82) | 146 (86) |

Continuous variables are expressed as the arithmetic mean and standard deviation (mean ± (SD)).

***Supplementary Table 7:*** *Urinary marker among different dietary groups*

| **Characteristic** | **High meat eater**<br>**N = 194,363** | **Low meat eater**<br>**N = 198,543** | **Poultry eater**<br>**N = 4,735** | **Pescatarian**<br>**N = 9,576** | **Vegetarian**<br>**N = 7,599** |
|---|---|---|---|---|---|
| Sodium (mmol/L) | 83 (46) | 73 (43) | 63 (41) | 67 (42) | 73 (48) |

| Characteristic | High meat eater N = 194,363 | Low meat eater N = 198,543 | Poultry eater N = 4,735 | Pescatarian N = 9,576 | Vegetarian N = 7,599 |
|---|---|---|---|---|---|
| Potassium (mmol/L) | 65 (34) | 62 (34) | 59 (35) | 59 (35) | 59 (35) |
| Creatinine (mmol/L) | 9.50 (5.88) | 8.35 (5.60) | 7.32 (5.46) | 7.15 (5.24) | 7.25 (5.43) |
| Microalbumin (mg/L) | 9 (58) | 7 (47) | 7 (61) | 6 (34) | 6 (45) |

Continuous variables are expressed as the arithmetic mean and standard deviation (mean ± (SD)).

***Supplementary Figure 2****: Subgroup analysis with hazard ratios (HR) and 95% confidence intervals (95% CI) between diet groups and the risk of chronic kidney disease (CKD) among people with normal kidney function and people having CKD Stage 1 or 2 at baseline. Age included as the underlying time variable. Model adjusted for sex, income and education level, waist circumference, BMI, physical activity and smoking status and stratified by ethnicity and T2DM diagnosis.*

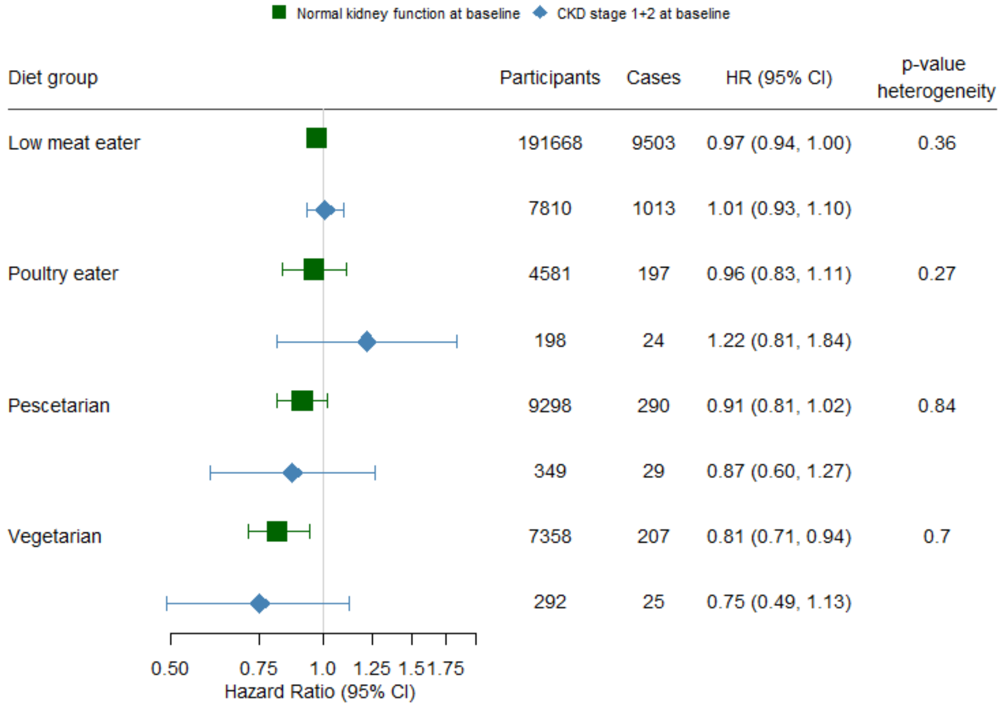

***Supplementary Figure 3****: Subgroup analysis with hazard ratios (HR) and 95% confidence intervals (95% CI) between diet groups and the risk of chronic kidney disease (CKD) among females and males. Age included as the underlying time variable.*

*Model adjusted for income and education level, waist circumference, BMI, physical activity and smoking status and stratified by ethnicity and T2DM diagnosis.*

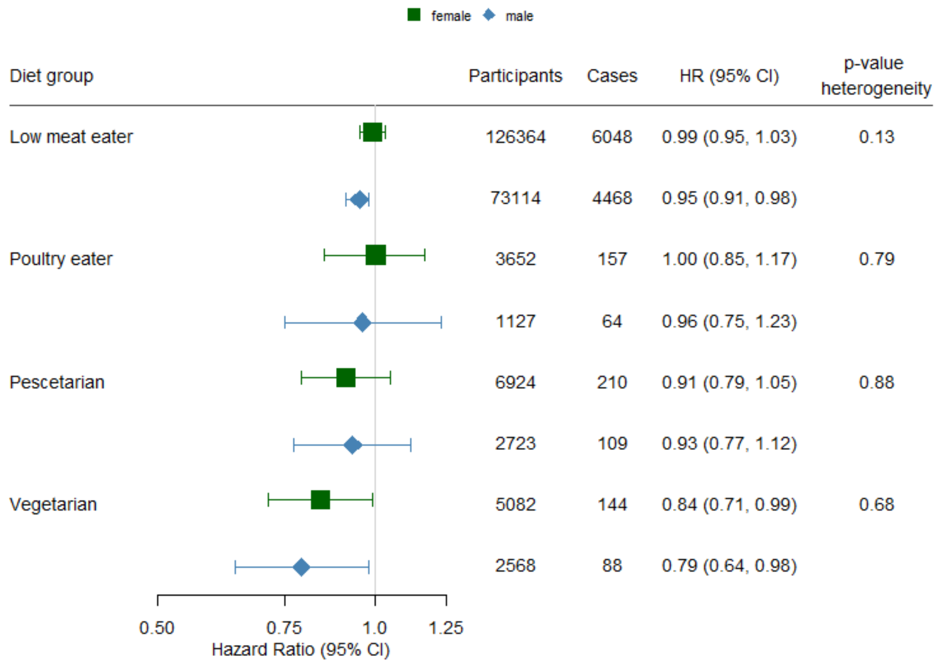

***Supplementary Figure 4*** *Subgroup analysis with hazard ratios (HR) and 95% confidence intervals (95% CI) between diet groups and the risk of chronic kidney disease (CKD) among people with low, medium and high polygenic risk. Age included as the underlying time variable. Model adjusted for sex, income and education level, waist circumference, BMI, physical activity and smoking status and stratified by ethnicity and T2DM diagnosis.*

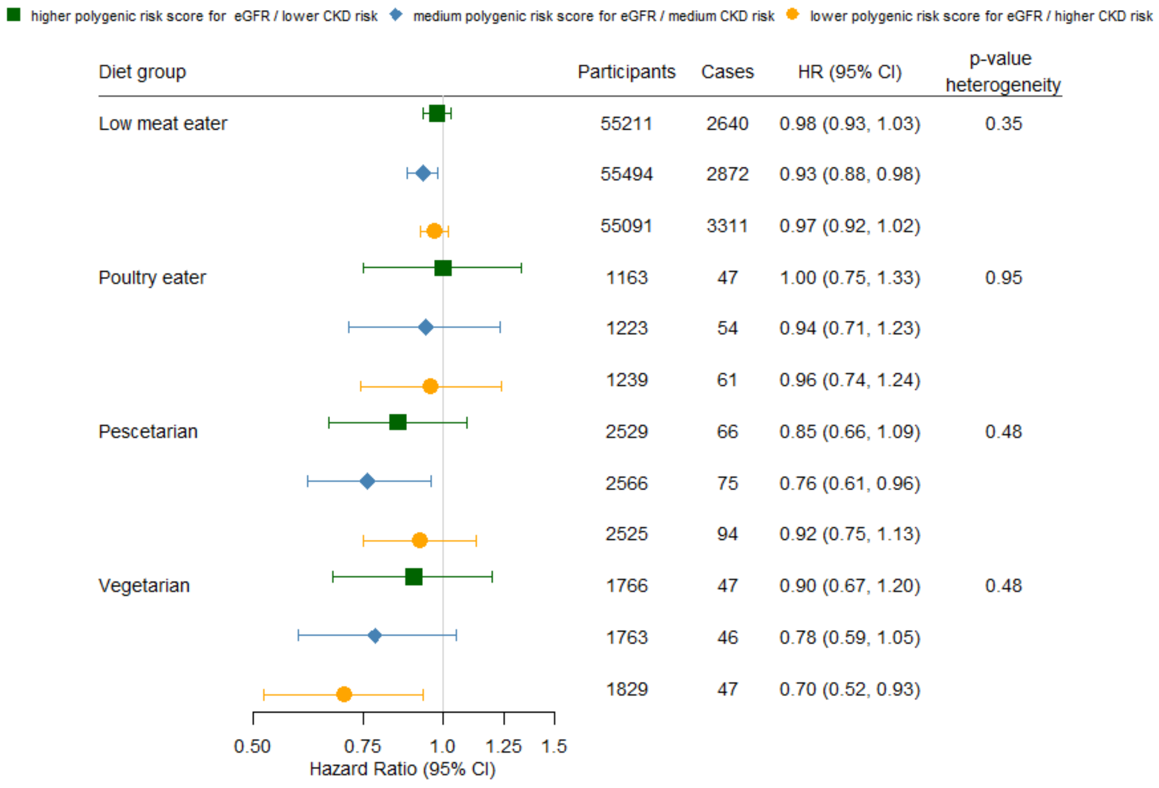

***Supplementary Figure 5:*** *Subgroup analysis with hazard ratios (HR) and 95% confidence intervals (95% CI) between diet groups and the risk of chronic kidney disease (CKD) among individuals with BMI lower or higher as 25. Age included as the underlying time variable. Model adjusted for sex, income and education level, waist circumference, physical activity and smoking status and stratified by ethnicity and T2DM diagnosis.*

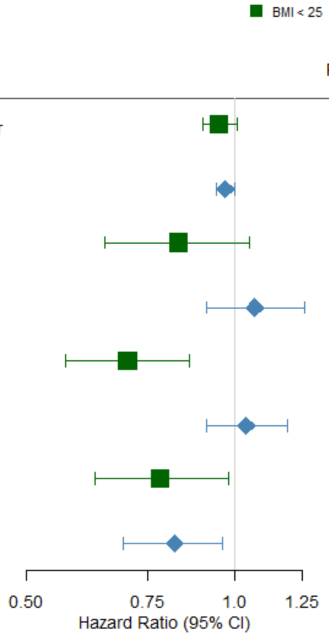

| Diet group | Participants | Cases | HR (95% CI) | p-value heterogeneity |
|---|---|---|---|---|
| Low meat eater | 72505 | 2472 | 0.95 (0.90, 1.01) | 0.59 |
| | 126973 | 8044 | 0.97 (0.94, 1.00) | |
| Poultry eater | 2480 | 71 | 0.83 (0.65, 1.05) | 0.08 |
| | 2299 | 150 | 1.07 (0.91, 1.26) | |
| Pescetarian | 5226 | 98 | 0.70 (0.57, 0.86) | 0.001 |
| | 4421 | 221 | 1.04 (0.91, 1.19) | |
| Vegetarian | 3898 | 83 | 0.78 (0.63, 0.98) | 0.77 |
| | 3752 | 149 | 0.82 (0.69, 0.96) | |

***Supplementary Figure 6:*** *Subgroup analysis with hazard ratios (HR) and 95% confidence intervals (95% CI) between diet groups and the risk of chronic kidney disease (CKD) among individuals low, medium and high education. Age included as the underlying time variable. Model adjusted for sex, income and education level, waist circumference, physical activity and smoking status and stratified by ethnicity and T2DM diagnosis.*

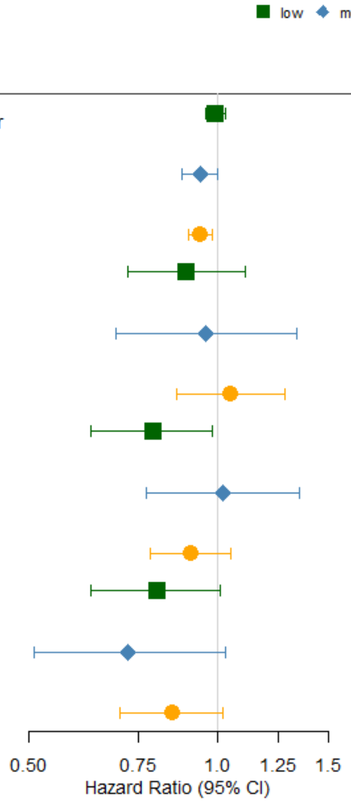

| Diet group | Participants | Cases | HR (95% CI) | p-value heterogeneity |
|---|---|---|---|---|
| Low meat eater | 66326 | 4768 | 0.99 (0.96, 1.03) | 0.14 |
| | 35023 | 1697 | 0.94 (0.88, 1.00) | |
| | 96249 | 3925 | 0.94 (0.90, 0.98) | |
| Poultry eater | 1412 | 81 | 0.89 (0.72, 1.11) | 0.56 |
| | 792 | 36 | 0.96 (0.69, 1.34) | |
| | 2506 | 100 | 1.05 (0.86, 1.28) | |
| Pescetarian | 1739 | 79 | 0.79 (0.63, 0.98) | 0.35 |
| | 1352 | 51 | 1.02 (0.77, 1.35) | |
| | 6490 | 181 | 0.91 (0.78, 1.05) | |
| Vegetarian | 1586 | 77 | 0.80 (0.63, 1.01) | 0.73 |
| | 1209 | 32 | 0.72 (0.51, 1.03) | |
| | 4753 | 121 | 0.85 (0.70, 1.02) | |

***Supplementary Figure 7****: Sensitivity analysis with hazard ratios (HR) and 95% confidence intervals (95% CI) between diet groups and the risk of chronic kidney disease (CKD). Age included as the underlying time variable. Basemodel adjusted for sex, income and education level, waist circumference, BMI, physical activity and smoking status and stratified by ethnicity and T2DM diagnosis. Basemodel including biomarker additionally adjusted for estimated glomerular filtration rate (eGFR) and urinary albumin to creatinine ratio (ACR).*

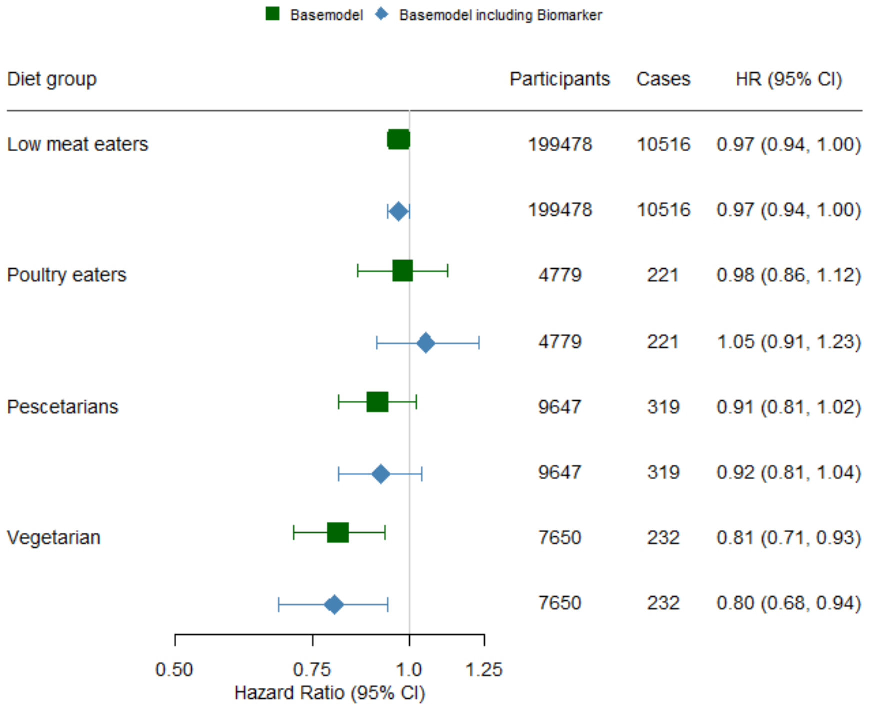

***Supplementary Figure 8:*** Sensitivity *analysis with hazard ratios (HR) and 95% confidence intervals (95% CI) between diet groups and the risk of chronic kidney disease (CKD). Age included as the underlying time variable. Basemodel adjusted for sex, income and education level, waist circumference, BMI, physical activity and smoking status and stratified by ethnicity and T2DM diagnosis. Basemodel including polygenic risk (PR) additionally adjusted for polygenic risk.*

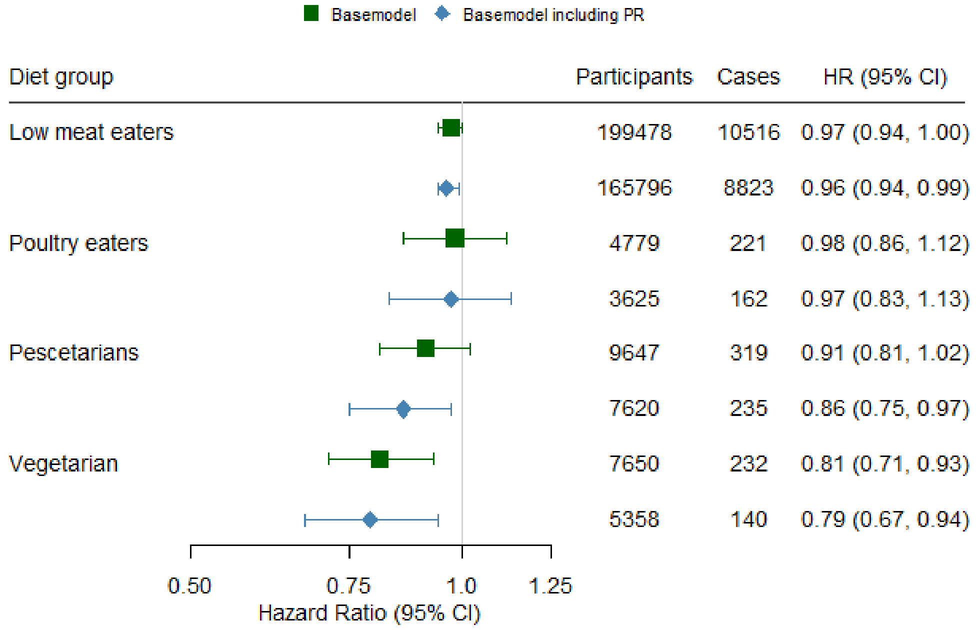
